# Beyond *R*_0_: Heterogeneity in secondary infections and probabilistic epidemic forecasting

**DOI:** 10.1101/2020.02.10.20021725

**Authors:** Laurent Hébert-Dufresne, Benjamin M. Althouse, Samuel V. Scarpino, Antoine Allard

## Abstract

The basic reproductive number — *R*_0_ — is one of the most common and most commonly misapplied numbers in public health. Although often used to compare outbreaks and forecast pandemic risk, this single number belies the complexity that two different pathogens can exhibit, even when they have the same *R*_0_ [1–3]. Here, we show how to predict outbreak size using estimates of the distribution of secondary infections, leveraging both its average *R*_0_ and the underlying heterogeneity. To do so, we reformulate and extend a classic result from random network theory [4] that relies on contact tracing data to simultaneously determine the first moment (*R*_0_) and the higher moments (representing the heterogeneity) in the distribution of secondary infections. Further, we show the different ways in which this framework can be implemented in the data-scarce reality of emerging pathogens. Lastly, we demonstrate that without data on the heterogeneity in secondary infections for emerging infectious diseases like COVID-19, the uncertainty in outbreak size ranges dramatically. Taken together, our work highlights the critical need for contact tracing during emerging infectious disease outbreaks and the need to look beyond *R*_0_ when predicting epidemic size.

## I. INTRODUCTION

In 1918, a typical individual infected with influenza transmitted the virus to between one and two of their social contacts [5], giving a value of the basic reproductive number – *R*_0_, the number of secondary infections in a completely susceptible population – between one and two. These are similar to values of *R*_0_ for the 2014 West Africa Ebola virus outbreak, but most individuals infected with Ebola virus gave rise to zero additional infections, while a few gave rise to more than 10 [6, 7]. Moreover, Ebola virus disease infected a tenth of one percent of the number of individuals believed to have been infected by the 1918 Influenza virus [8, 9]. While improvements in healthcare and public health measures, as well as changes in human behavior, partially explain the massive discrepancy between Ebola virus disease in 2014 and influenza in 1918 [10], there is another critical difference between these two diseases: heterogeneity in the number secondary cases resulting from a single infected individual. Here, we demonstrate analytically that quantifying the variability in the number of secondary infections is critically important for quantifying the transmission risk of novel pathogens.

The basic reproduction number of an epidemic, *R*_0_, is the expected number of secondary cases (note, we use the word “case” in a generic sense to represent any infection, even if too mild to meet the clinical case definition) produced by a primary case over the course of their infectious period in a completely susceptible population [11]. It is a simple metric that is commonly used to describe and compare the transmissibilty of emerging and endemic pathogens [12]. If *R*_0_ = 2, one case turns to two, on average, and two turn to four as the epidemic grows. Conversely, the epidemic will die out if *R*_0_ < 1.

Almost 100 years ago, work from Kermack and McKendrick [13–15] first demonstrated how to estimate the final size of an epidemic. Specifically, they considered a scenario such that: (*i*) the disease results in complete immunity or death, (*ii*) all individuals are equally susceptible, (*iii*) the disease is transmitted in a closed population, (*iv*) contacts occur according to the law of mass-action, (*v*) and the population is large enough to justify a deterministic analysis. Under these assumptions, Kermack and McKendrick show that an epidemic with a given *R*_0_ will infect a fixed fraction *R*(∞) of the susceptible population by solving

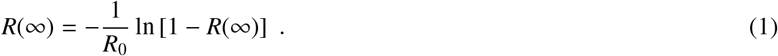

This solution describes a final outbreak size equal to 0 when *R*_0_ ≤ 1 and increasing roughly as 1 − exp(*R*_0_) when *R*_0_ > 1. Therefore, a larger *R*_0_ leads to a larger outbreak which infects the entire population in the limit *R*_0_→ ∞. This direct relationship between *R*_0_ and the final epidemic size is at the core of the conventional wisdom that a larger *R*_0_ will cause a larger outbreak. Unfortunately, the equation relating *R*_0_ to final outbreak size from Kermack and McKendrick is only valid when all the above assumptions hold, which is rarely the case in practice.

As a result, relying on *R*_0_ alone is often misleading when comparing different pathogens or outbreaks of the same pathogen in different settings [1–3]. This is especially critical considering that many outbreaks are not shaped by the “average” individuals but rather by a minority of super-spreading events [1, 16]. To more fully quantify how heterogeneity in the number of secondary infections affects outbreak size, we turn towards network epidemiology and derive an equation for the total number of infected individuals using all moments of the distribution of secondary infections.

## II. RANDOM NETWORK ANALYSIS

Random network theory allows us to relax some of assumptions made by Kermack and McKendrick, mainly to account for heterogeneity and stochasticity in the number of secondary infections caused by a given individual. We first follow the analysis of Ref. [4] and define

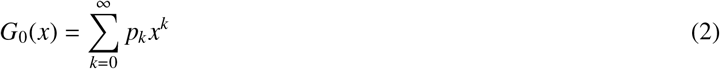

as the probability generating function (PGF) of the distribution of the number on contacts *p*_*k*_ individuals have (the *degree* distribution). When following a random contact (an *edge*), we define the *excess* degree as the number of other edges around that node reached via one of its edges. Because an edge is *k* times more likely to reach a node of degree *k* than a node of degree 1, the excess degree distribution is generated by

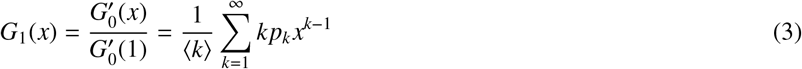

where ⟨*k*⟩ is the average degree and acts as a normalisation constant, and 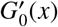 denotes the derivative of *G*_0_(*x*) with respect to *x*. We now assume that the network in question is the network of all edges that *would* transmit a disease if given the chance. Consequently, *G*_1_(*x*) generates the number of secondary infections that individual nodes would cause if infected. And, if we infect a random node as the patient zero, its entire connected component (a maximal subset of nodes between which paths exists between all pairs of nodes) will be infected. To calculate the largest possible epidemic, we thus look for the size of the giant connected component (GCC).

To calculate the size of the GCC, we first look for the probability *u* that following a random edge leads to a node *not* part of the GCC. For that node to not be part of the GCC, all of its excess edges must also not lead to the GCC. This simple observation leads to the self-consistent equation

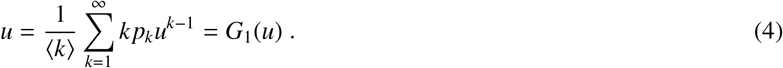

The size of the GCC is a fraction of the full population *N* that we will denote *R*(∞) because it corresponds to the potential, macroscopic, outbreak size. Noting that a node of degree *k* is *not* in the GCC with probability *u*^*k*^, *R*(∞) can be calculated by counting all nodes except those with no edges leading to the GCC,

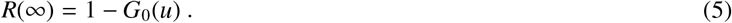

When dealing with data on the distribution of secondary infections, *G*_0_(*x*) has one degree of freedom remaining, *p*_0_, which we set by assuring that the number of infections caused by patient zero is smaller or equal to *R*_0_ but not greater (see Methods). The resulting solution for *R*(∞) is exact in the limit of infinite population size.

## III. RESULTS

The network approach naturally accounts for heterogeneity, meaning that some individuals will cause more infections than others. The network approach also accounts for stochasticity explicitly: Even with *R*_0_ > 1, there is a probability 1− *R*(∞) that patient zero lies outside of the giant outbreak and therefore only leads to a small outbreak that does not invade the population. However, the analysis in terms of PGFs is obviously more involved than simply assuming mass-action mixing and solving Eq. (1). In fact, the PGF *G*_0_(*x*) requires a full distribution of secondary cases per primary case, which will in practice involve the specification of a high-order polynomial.

To clarify and potentially simplify the approach, we propose to reformulate the classic network model in terms of the cumulant generating function (CGF) of secondary cases. The CGF *K*(*y*) of a random variable *X* can be written as *K*(*y*) = *κ*_*n*_*y*^*n*^/*n*! where *κ*_*n*_ are the cumulants of the distribution of secondary infections. These are useful because the cumulants are easier to interpret, i.e., *κ*_1_ is simply the average number of secondary cases *R*_0_, *κ*_2_ is the variance, *κ*_3_ is related to the skewness and *κ*_4_ to the kurtosis of the full distribution, etc. By definition, a PGF *G*(*x*) of a random variable is linked to *K*(*y*) through *G*(*x*) = exp [*K* (ln *x*)]. Therefore, we can replace the PGF *G*_1_(*x*) for the distribution of secondary infections by a function in terms of the cumulants of that distribution.

### A. Analysis of cumulants and derivation of Kermack-McKendrick

We can easily derive Kermack and McKendrick’s result from this framework since their solution assumes a well-mixed population, which corresponds to a Poisson distribution of secondary infections. We first re-write *G*_1_(*x*) in terms of the cumulants *κ*_*n*_ as

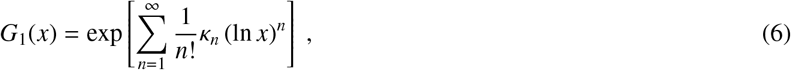

which is a particular convenient representation for a Poisson distribution because its cumulants *κ*_*n*_ = *R*_0_ for all *n* > 0. Moreover, since *G*_0_(*x*) = *G*_1_(*x*) in the Poisson case, the final outbreak size of the Kermack-McKendrick analysis will be set by *u*_KM_ = *G*_1_(*u*_KM_), or

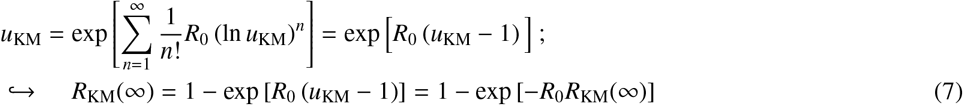

Taking the logarithm of the exponential term from this last equation yields Kermack and McKendrick’s formula. The solution to *u* = *G*_1_(*u*) gives the probability that every infection caused by patient zero fails to generate an epidemic. For more general distributions, it is useful to rewrite Eq. (6) as

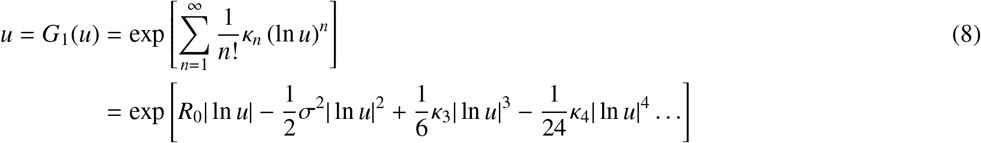

to highlight its alternating nature because ln *u* is negative (*u* is a probability) such that its *n*-th power is positive when *n* is even and negative when *n* is odd.

The alternating sign of contribution from high-order moments in Eq. (8) can be interpreted as follows. A disease needs a high average number of secondary infections (high *κ*_1_ = *R*_0_) to spread, but given that average, a disease with small variance in secondary infections will spread much more reliably and be less likely to stochastically die out. Given a variance, a disease with high skewness (i.e., with positive deviation contributing to most of the variance) will be more stable than a disease with negative skewness (i.e. with most deviations being towards small secondary infections). Given a skewness, a disease will be more stable if it has frequent small positive deviations rather than infrequent large deviations — hence a smaller kurtosis — as stochastic die out could easily occur before any of those large infrequent deviations occur.

Our re-interpretation already highlights a striking result: Higher moments of the distribution of secondary cases can lead a disease with a lower *R*_0_ to invade more easily a population and to reach a larger final outbreak size than a disease with a higher *R*_0_. Taking into account the contribution of these higher moments also yields better estimates for the final size of outbreaks, as we now show.

### B. Comparison of estimators to empirical data

We now compare the final outbreak size estimates from Eq. (1) (Kermack and McKendrick) to estimates from Eq. (5) (network model) with a negative binomial offspring distribution (see Methods and Table I). As predicted, Fig. 1 (left) shows that the Kermack and McKendrick formulation consistently and significantly over-predicts the outbreak size across six different pathogens where we could find confidence interval estimates for *R*_0_ and for the negative binomial over-dispersion parameter (*k*). Our approach produces estimates of the total outbreak size, which are consistent with outbreaks where no vaccine was available (smallpox in unvaccinated populations, the 1918 influenza pandemic, and school children prior to the availability of the 2009 H1N1 vaccine). Clearly, once interventions are put in place and/or substantial behavioral change occurs, all methods that do not account for these effects will over-estimate the total outbreak size [30]. Nevertheless, our approach provides a much more reasoned estimate of the total risk to any given population, and predictions very close to the most recent seropositivity estimates for the COVID-19 outbreak in a German Municipality [28] and in obstetrical patients presenting for delivery [29], as well as for SARS among wild animal handlers (other smaller estimates correspond to health-care workers) [19].

**FIG. 1.**
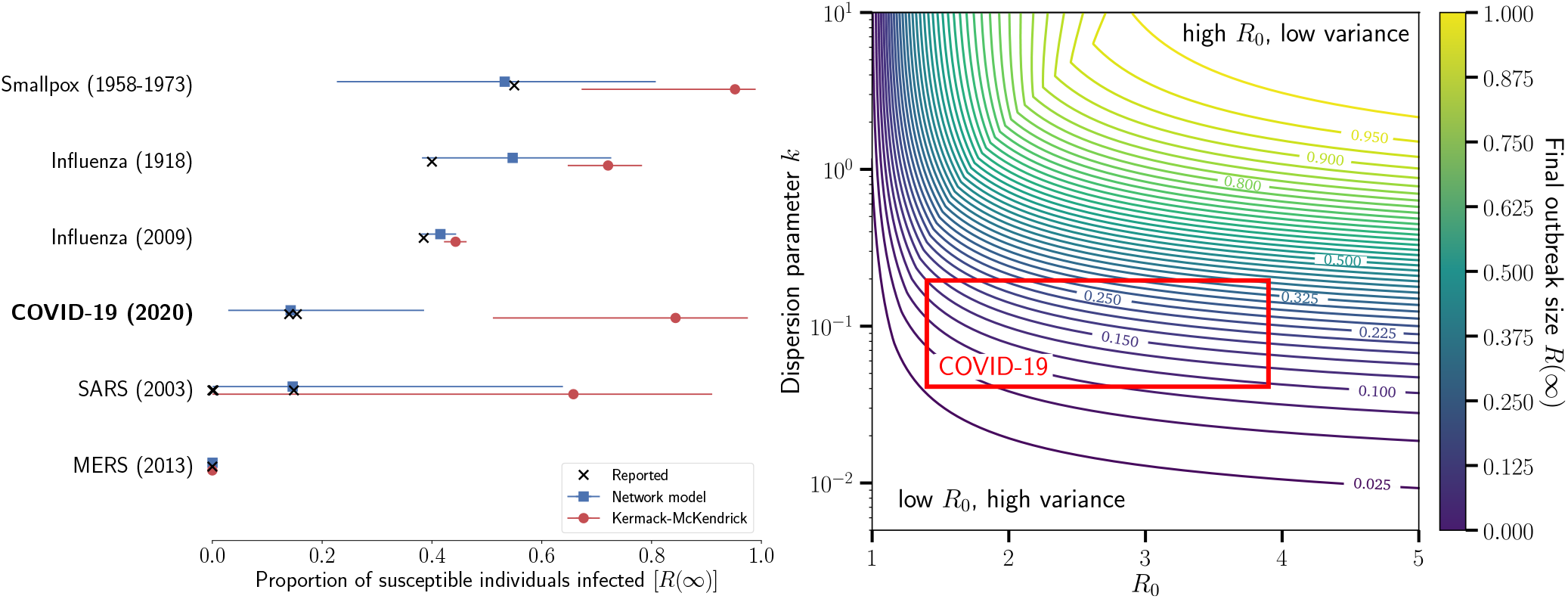
(left) Using published estimates of *R*_0_ and the dispersion parameter, *k*, we estimated the total outbreak size for six different diseases. The confidence intervals span the range of uncertainty reported for *R*_0_ and *k*. The black markers show reported total outbreak sizes (total proportion of susceptible individuals infected) for each disease. For influenza, we report the estimated proportion of school-aged children infected. The blue squares show the estimated proportion infected obtained with Eq. (5). The red circles are the estimated proportion infected using the method developed by Kermack and McKendrick, i.e., Eq. (1). (right) Final size of outbreaks with different *R*_0_ and distributions of secondary cases. We use a negative binomial distribution of secondary cases and scan a realistic range of parameters. The range of parameters corresponding to current estimates for COVID-19 is highlighted by a red box. Most importantly, with fixed average, the dispersion parameter is inversely proportional to the variance of the underlying distribution of second cases. See Table I and https://github.com/Emergent-Epidemics/beyond_R0 for additional information and to access the data.

**TABLE I.** Estimates for *R*_0_ and for the negative binomial distribution dispersion parameter, *k*, used on Fig. 1 (§ and ★ respectively denote 95% and 90% confidence intervals). Proportion of susceptible individuals infected as reported in either the literature or by the US CDC. For SARS, the proportion of infected was taken from serosurveys among wild animal handlers (0.15) and among health-care workers (<0.01) [19]. For influenza (2009), we took data on school-aged children. For COVID-19, the proportion of infected was taken from a serosurvey in the municipality of Gangelt, Germany [28] and from universal testing in all obstetrical patients presenting for delivery at two hospitals [29]. Note that the estimates the proportion of infected individuals, for *R*_0_ and for *k* were not necessarily inferred from the same populations. Such information is rarely, if ever, available for a same outbreak, unfortunately.

| Disease | Location | Year | Prop. Infect. | $R_0$ | $k$ | Ref. |
| --- | --- | --- | --- | --- | --- | --- |
| MERS | Global | 2013 | 0.00 | 0.47 (0.29–0.80)<sup>§</sup> | 0.26 (0.09–1.24)<sup>§</sup> | [17, 18] |
| SARS | Global | 2003 | 0.00–0.15 | 1.63 (0.54–2.65)<sup>★</sup> | 0.16 (0.11–0.64)<sup>★</sup> | [1, 19, 20] |
| Smallpox | Europe | 1958–1973 | 0.55 | 3.19 (1.66–4.62)<sup>★</sup> | 0.37 (0.26–0.69)<sup>★</sup> | [1, 21] |
| Influenza | Baltimore (USA) | 1918 | 0.40 | 1.77 (1.61–1.95)<sup>§</sup> | 0.94 (0.59–1.72)<sup>§</sup> | [22, 23] |
| Influenza | Italy | 2009 | 0.39 | 1.321 (1.299–1.343)<sup>§</sup> | 8.092 (5.170–11.794)<sup>§</sup> | [24, 25] |
| COVID-19 | Global | 2020 | 0.14, 0.154 | 2.5 (1.4–12)<sup>§</sup> | 0.1 (0.04–0.2)<sup>§</sup> | [26–29] |

## IV. DISCUSSION

From re-emerging pathogens like yellow fever and measles to emerging threats like Middle East Respiratory Syndrome coronavirus and Ebola, the World Health Organization monitored 119 different infectious disease outbreaks in 2019 alone [31]. For each of these outbreaks, predicting both the epidemic potential and the most likely number of cases is critically important for efficient and effective responses. This need for rapid situational awareness is why *R*_0_ is so widely used in public health. However, our main analysis shows that not only is *R*_0_ insufficient in fully determining the final size of an outbreak, but having a larger outbreak with a lower *R*_0_ is relatively easy considering the randomness associated with most transmission events and the heterogeneity of physical contacts. To address the need for rapid quantification of risk, while acknowledging the shortcomings of *R*_0_, we use network science methods to derive both the probability of an epidemic and its final size.

These results are not without important caveats. Specifically, we must remember that distributions of secondary cases, just like *R*_0_ itself, are just as much a product of a pathogen as of the population in which it spreads. For example, aspects of the social contact network [32], metapopulation structure [33], human mobility [34], adaptive behavior [35], and even other pathogens [36, 37], all interact to cause complex patterns of disease emergence, spread, and persistence. Therefore, great care must be taken when using any of these tools to compare outbreaks or to inform current events with past data.

Figure 1 (left) only used a handful of known outbreaks to validate the different approaches because data on secondary cases are rare. In practice, three types of data could potentially be used in real time to improve predictions by considering secondary case heterogeneity. First, contact tracing data whose objective is to identify people who may have come into contact with an infectious individual. While mostly a preventive measure to identify cases before complications, it directly informs us about potential secondary cases caused by a single individual, and therefore provides us with an estimate for *G*_1_(*x*). Both for generating accurate predictions of epidemic risk and controlling the outbreak, it is vital to begin contact tracing before numerous transmission chains become widely distributed across space [38, 39].

Second, viral genome sequences provide information on both the timing of the outbreak [40] and structure of secondary cases [41]. For example, methods exist to reconstruct transmission trees for sampled sequences using simple mutational models to construct a likelihood for a specific transmission tree [42, 43] and translate coalescent rates into key epidemiological parameters [44, 45]. Despite the potential for genome sequencing to revolutionize outbreak response, the global public health community still struggles to coordinate data sharing across international borders, between academic researchers, and with private companies [46–48].

Third, early incidence data can be leveraged to infer parameters of the secondary case distribution through comparison with simulations. Comparing the output of agent-based simulations with reported incidence can be used to effectively sample a joint posterior distribution over *R*_0_ and dispersion parameter *k*. This approach was used by most studies referenced in Table I. Most importantly, these simulations need not be run over long periods of time to predict final outbreak size. Instead, they only need to be run over enough early data to infer the parameter estimates that are then fed into our network model to compute the final outbreak size.

As for COVID-19, Fig. 1(right) shows how the width of the confidence interval on our prediction for the final outbreak size mostly stems from uncertainty in the heterogeneity of secondary infections; i.e., the dispersion parameter *k*. With limited heterogeneity, our predictions would have been closer to classic mass-action forecasts and the current pandemic of COVID-19 would likely have been a consequence of not only *R*_0_, but of the homogeneity of secondary infections: each new cases steadily leading to additional infections. Thankfully, with recent large estimates for its heterogeneity, the observed transmission could be mostly maintained by so-called “super-spreading events”, which could be easier to manage with contact tracing, screening and infection control [49].

In conclusion, we reiterate that when accounting for the full distribution of secondary cases caused by an infected individual, there is no direct relationship between *R*_0_ and the size of an outbreak. We also stress that both *R*_0_ and the full secondary case distribution are not properties of the disease itself, but are instead set by properties of the pathogen, the host population and the context of the outbreak. Nevertheless, we provide a straightforward methodology for translating estimates of transmission heterogeneity into epidemic forecasts. Altogether, predicting outbreak size based on early data is an incredibly complex challenge but one that is increasingly within reach due to new mathematical analyses and faster communication of public health data.

## Data Availability

Non applicable

https://github.com/Emergent-Epidemics/beyond_R0

## ACKNOWLEDGMENTS

L.H.-D. acknowledges support from the National Institutes of Health 1P20 GM125498-01 Centers of Biomedical Research Excellence Award. B.M.A. is supported by Bill and Melinda Gates through the Global Good Fund. S.V.S. is supported by startup funds provided by Northeastern University. A.A. acknowledges financial support from the Sentinelle Nord initiative of the Canada First Research Excellence Fund and from the Natural Sciences and Engineering Research Council of Canada (project 2019-05183).

## METHODS

The results presented from our network model assume the number of secondary infections to be distributed according to a negative binomial distribution parametrized by its average *R*_0_ and dispersion *k* [1]. Its probability generating function (PGF) is

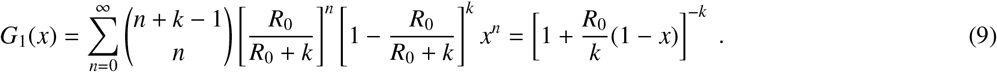

The network theory formalism presented in the main text requires the specification of the PGF *G*_0_(*x*) whose related to *G*_1_(*x*) via

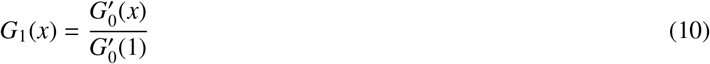

where the prime (′) denotes the first derivative with respect to *x*. Specifying *G*_1_(*x*) therefore fixes *G*_0_(*x*) up to a constant and to a multiplicative factor. Without loss of generality, we set

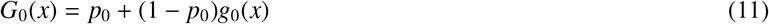

with 0 ≤ *p*_0_ ≤ 1, *g*_0_(0) = 0 and *g*_1_(1) = 1. Equation (10) becomes

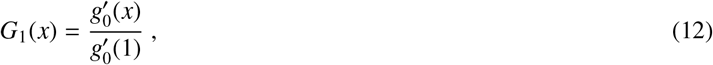

from which we compute

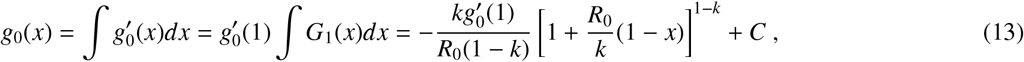

with *k* ≠ 1, and where *C* and 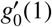 are fixed by imposing *g*_0_(0) = 0 and *g*_1_(1) = 1. Rearranging the terms, we find that

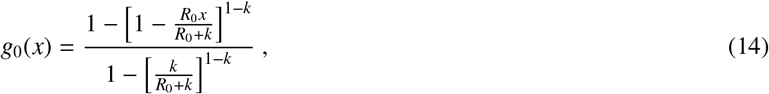

from which we finally obtain

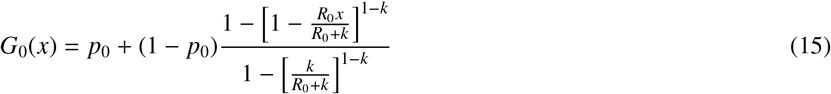

with *k* ≠ 1. The case *k* = 1 must be treated separately and yields

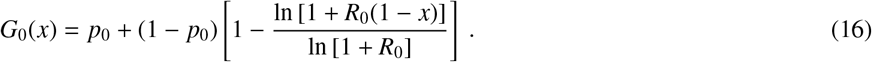

From Eqs. (15) and (16), we find that the average number of secondary infections caused by the patient zero is

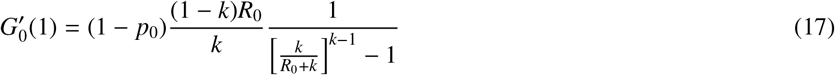

if *k* ≠1, and

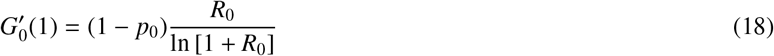

if *k* = 1. The average number of secondary infections caused by patient zero can therefore be greater or smaller than *R*_0_. Since patient zero should not be expected to create *more* secondary cases than the next generation of infections, we set the value of *p*_0_ ∈ [0, 1] such that 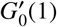 is as close as possible to *R*_0_ whenever 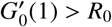.

A large-scale epidemic is predicted by this framework [4] if

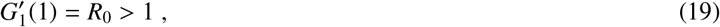

as in the analysis by Kermack and McKendrick [13–15]. Its size, *R*(∞), is computed with *G*_0_(*x*) as

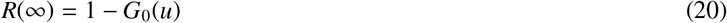

where *u* is the solution of

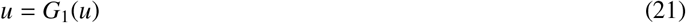

which we solve using the relaxation method [50] with an initial condition randomly chosen in the open interval (0, 1).

